# Night Shift Work Increases the Risk of Asthma

**DOI:** 10.1101/2020.04.22.20074369

**Authors:** RJ Maidstone, J Turner, C Vetter, HS Dashti, R Saxena, FAJL Scheer, SA Shea, SD Kyle, DA Lawlor, ASI Loudon, JF Blaikley, MK Rutter, DW Ray, HJ Durrington

## Abstract

Shift work causes misalignment between our internal clock and daily behavioural cycles and is associated with metabolic disorders and cancer. Here, we describe the relationship between shift work and prevalent asthma in >280,000 UK Biobank participants. Compared to day workers, ‘permanent’ night shift workers had a higher likelihood of moderate/severe asthma (odds ratio (OR) 1.36 (1.03-1.8)) and all asthma (OR 1.23 (1.03-1.46) after adjustment for known major confounders). The public health implications of this finding are far-reaching due to the high prevalence and co-occurrence of both asthma and shift work.

## Introduction

Most human biological processes are regulated by an internal circadian timing system to optimally prepare physiological functions for the anticipated daily environmental and behavioural cycles. Cyclical light/dark environmental cues, mealtimes and physical activity can serve as *Zeitgebers* for the circadian timing system. The development of artificial light has allowed extension of the active period of humans into the night, and through the night for night shift workers. This imbalance between our internal clock and the environment results in circadian misalignment (1). Shift work is a notable example of circadian misalignment, is invariably associated with sleep disruption and with increased risk of prevalent, chronic diseases including obesity (2), metabolic syndrome (3), diabetes (4), cardiovascular disease (5), and cancers (6, 7). There is evidence of causal relationships between circadian misalignment and the development of diabetes, obesity, metabolic syndrome (8) and cardiovascular disease (9). In mice, experimentally induced circadian disruption (by altering light/dark cycle, to simulate rotating shift work patterns) affects the innate immune system and inflammation (10).

Approximately 20% of the working population in industrialized countries work permanent or rotating night shifts (11), exposing this large population to the risk of circadian misalignment-driven disease; making this is an important area of investigation, and an emerging public health emergency. Analysis of the impact of shift work on chronic inflammatory diseases is lacking.

Asthma is a very common, chronic inflammatory disease of the airways; affecting 339 million people worldwide (12) and costing the UK public sector £1.1 billion (13) ($80 billion in the US each year (14)). Intriguingly, asthma displays marked time of day variations in symptoms (wheeze and whistling) (15), airway calibre (16), and in the underpinning inflammatory pathways (17). The physiological diurnal variation in airway calibre is under direct circadian control, independent of external, environmental cues such as light/dark and fasting or feeding (18). In asthma, it appears that the physiological diurnal variation in airway calibre is amplified, suggesting coupling between the internal body clock and pathogenic processes. This raises the possibility that misalignment between the internal body clock and the environment, such as that induced by night shift work, would impact on asthma risk. Indeed a correlation between shift work and work-related asthma was found in a study of 544 individuals working in a cabling manufacturing plant (19). Therefore, we investigated the association between shift work and asthma in a much larger dataset from the UK Biobank (20) in which we could also adjust for numerous major confounding factors such as smoking history, race and ethnicity, socio-economic status, physical activity, and BMI.

We hypothesised that when compared to day workers, both current and past shift work, especially involving nights, would be associated with a higher prevalence of asthma.

We also investigated whether chronotype is associated with the risk of asthma in shift workers. Chronotype is the phenotypic expression of the internal circadian timing system and shows substantial variation in the general population with women typically being more morning types than men, and adolescents showing later circadian phenotypes than younger children and adults (21, 22). Chronotype can affect how an individual adapts to shift work; earlier chronotypes experience shortened sleep duration and increased sleep disturbance during night shifts, whereas late chronotypes show similar disruption when working early shifts (23). Matching shift work patterns to chronotype can improve sleep quality and well-being (24).

Lastly, we investigated the intersection between genetic risk of asthma, and shift work exposure. Asthma risk was captured using a genetic risk score (GRS); sum of genetic variants with weighted effect sizes (25). If asthma GRS affects the health impact of shift work exposure this may provide an employment screening opportunity in the future.

## Results

### Demographics of Participants

UK Biobank recruited 502,540 participants (5% of those invited) aged 40 to 69 years who were registered with the National Health Service (NHS) and lived within reasonable traveling distance of 22 assessment centers across the UK between 2007 and 2010 (26). At the baseline visit, participants completed questionnaires on lifestyle, medical history, occupation and work hours; trained health professionals asked further details about medical conditons, health status and medications. The selection of participants analysed in all comparisons are detailed in a STROBE diagram (**Supplementary Figure 1**).

Analysis of shift work was restricted to participants in paid employment or who were self-employed at baseline (N=286,825, age range 37-72 years) (4); we did not exclude any individuals based on other diagnoses. The demographics of this group are shown in **Table 1**. Of these, 83% were day workers and 17% worked shifts of which 51% included night shifts. Compared to day workers, shift workers were more likely to be male, lived in more deprived neighbourhoods (Townsend area deprivation Index), more likely to live in an urban area and more likely to be smokers. Shift workers drank less alcohol, reported shorter sleep duration and longer weekly working hours. Night shift workers were more likely evening chronotypes compared to those working days. Shift workers were more likely of non-European ancestry, and to be in jobs linked to occupational asthma or to jobs that require a medical examination. Compared to day workers, shift workers were more likely to have a diagnosis of gastro-oesophageal reflux, chronic obstructive pulmonary disease (COPD)/emphysema, higher cholesterol and hypertension.

**Table 1:** Clinical characteristics by current shift work exposure (N = 286,825)

|  | Current work schedule |  |  |  |
| --- | --- | --- | --- | --- |
|  | Day workers | Shift work, but never or rarely night shifts | Irregular shift work including nights | Permanent night shift work |
| N | 236,897 | 24,560 | 18,226 | 7,142 |
| Age (years) | 52.90 (7.13) | 52.48 (7.08) | 51.08 (6.87) | 51.45 (6.91) |
| Sex (% male) | 46.58 | 47.51 | 62.43 | 61.43 |
| BMI (kg/m<sup>2</sup>) | 27.09 (4.65) | 27.79 (4.99) | 28.21 (4.91) | 28.51 (4.88) |
| **Smoker (%)** |  |  |  |  |
| Never | 58.10 | 53.66 | 52.82 | 51.99 |
| Previous | 31.91 | 32.11 | 30.52 | 30.03 |
| Current | 9.75 | 13.88 | 16.19 | 17.67 |
| Smoking pack-years | 20.07 (16.07) | 22.92 (17.49) | 24.31 (17.77) | 25.70 (18.38) |
| Daily alcohol intake (%) | 20.48 | 16.89 | 15.98 | 10.21 |
| Sleep Duration (h) | 7.05 (1.03) | 6.95 (1.22) | 6.85 (1.30) | 6.67 (1.52) |
| Morning Chronotype (%) | 23.33 | 25.49 | 22.85 | 19.24 |
| Evening Chronotype (%) | 8.02 | 7.87 | 9.83 | 16.90 |
| **Ethnicity (%)** |  |  |  |  |
| White British | 88.47 | 83.30 | 79.87 | 80.99 |
| White Other | 6.45 | 7.07 | 7.03 | 6.01 |
| Mixed | 0.65 | 0.90 | 0.97 | 0.87 |
| Asian | 1.72 | 3.58 | 3.84 | 3.39 |
| Black | 1.40 | 2.69 | 4.93 | 5.47 |
| Chinese | 0.34 | 0.48 | 0.46 | 0.67 |
| Other | 0.09 | 0.13 | 0.10 | 0.14 |
| Weekly work hours | 34.24 (13.19) | 34.97 (13.21) | 39.29 (14.55) | 39.59 (13.73) |
| Job Asthma Risk (%) | 7.59 | 7.18 | 8.11 | 7.74 |
| Job Medical Required (%) | 2.27 | 2.52 | 4.14 | 3.68 |
| Single Occupancy (%) | 15.64 | 18.78 | 18.71 | 18.42 |
| Urban area (%) | 85.98 | 89.59 | 89.33 | 90.97 |
| Townsend Index | -2.24 (-3.70 to 0.19) | -1.31 (-3.18 to 1.61) | -1.24 (-3.17 to 1.82) | -1.04 (-3.02 to 2.07) |
| Maternal Smoking (%) | 26.59 | 28.88 | 29.23 | 30.75 |
| Breastfed as baby (%) | 56.12 | 54.27 | 54.16 | 51.51 |
| Birth Weight (kg) | 3.33 (0.63) | 3.31 (0.68) | 3.35 (0.67) | 3.31 (0.71) |
| Hypertension (%) | 19.75 | 21.58 | 21.64 | 22.81 |
| High Cholesterol (%) | 7.88 | 8.55 | 8.54 | 9.27 |
| Sleep Apnoea (%) | 0.28 | 0.30 | 0.42 | 0.27 |
| Chronic Obstructive Pulmonary Disease(COPD) /Emphysema/Chronic Bronchitis (%) | 0.81 | 1.26 | 1.27 | 1.23 |
| Bronchiectasis (%) | 0.14 | 0.12 | 0.03 | 0.14 |
| Interstitial Lung Disease (%) | 0.02 | 0.01 | 0.03 | 0.01 |
| Other Respiratory Problems (%) | 0.12 | 0.17 | 0.16 | 0.07 |
| Gastro-Oesophageal Reflux (%) | 3.19 | 3.65 | 3.84 | 4.16 |

Data are mean (SD), median (IQR) or percentages. Positive values of the Townsend index indicate high material deprivation, negative values indicate relative affluence. The diagnosis of conditions (hypertension, high cholesterol, sleep apnoea, COPD/emphysema/chronic bronchitis, bronchiectasis,
interstitial lung disease, other respiratory problems and gastro-oesophageal reflux) came from participants self-reporting a doctor diagnosis.

### Cases of Asthma

Cases of asthma were defined by including all participants with self-reported doctor-diagnosed asthma at baseline who were also receiving any asthma medication (27). Using these criteria, we identified 14,238 (5.3%) cases, of which 4,783 (1.9%) had moderate-severe asthma (defined as having doctor diagnosed asthma at baseline and currently taking medication in accordance with step 3-5 of the British Thoracic Society guidance for the treatment of asthma) (27). We excluded from our analyses: participants with doctor-diagnosed asthma who did not report taking asthma medication as well as those participants reporting taking asthma medication who did not have doctor-diagnosed asthma (N=20,151). For analysis of moderate-severe asthma we further excluded those not on medication for moderate-severe asthma (listed in methods section; N = 9,455). Initially, we focussed on those with moderate-severe asthma, since these individuals were more likely to have active asthma requiring regular disease-modifying treatment, so reducing the risk of misdiagnosis.

In an age- and sex-adjusted model, there were higher odds of having moderate-severe asthma in shift workers who never or rarely undertook night shifts (OR 1.12 (95% CI: 1.02-1.24) and in those on permanent night shifts (OR 1.21 (1.02-1.44)) when compared to day workers, **Figure 1**. After further adjusting for smoking status and pack years, alcohol status and intake, ethnicity, social deprivation, physical activity, BMI, chronotype, length of working week, job asthma risk and job medical required (model 2), associations attenuated in shift workers who never or rarely undertook night shifts (OR 1.17 (0.98-1.38)) and slightly increased in permanent night shift workers (OR 1.36 (1.03-1.8)). Further adjustment for sleep duration had no additional effects on the estimates (model 3).

**Figure 1:**
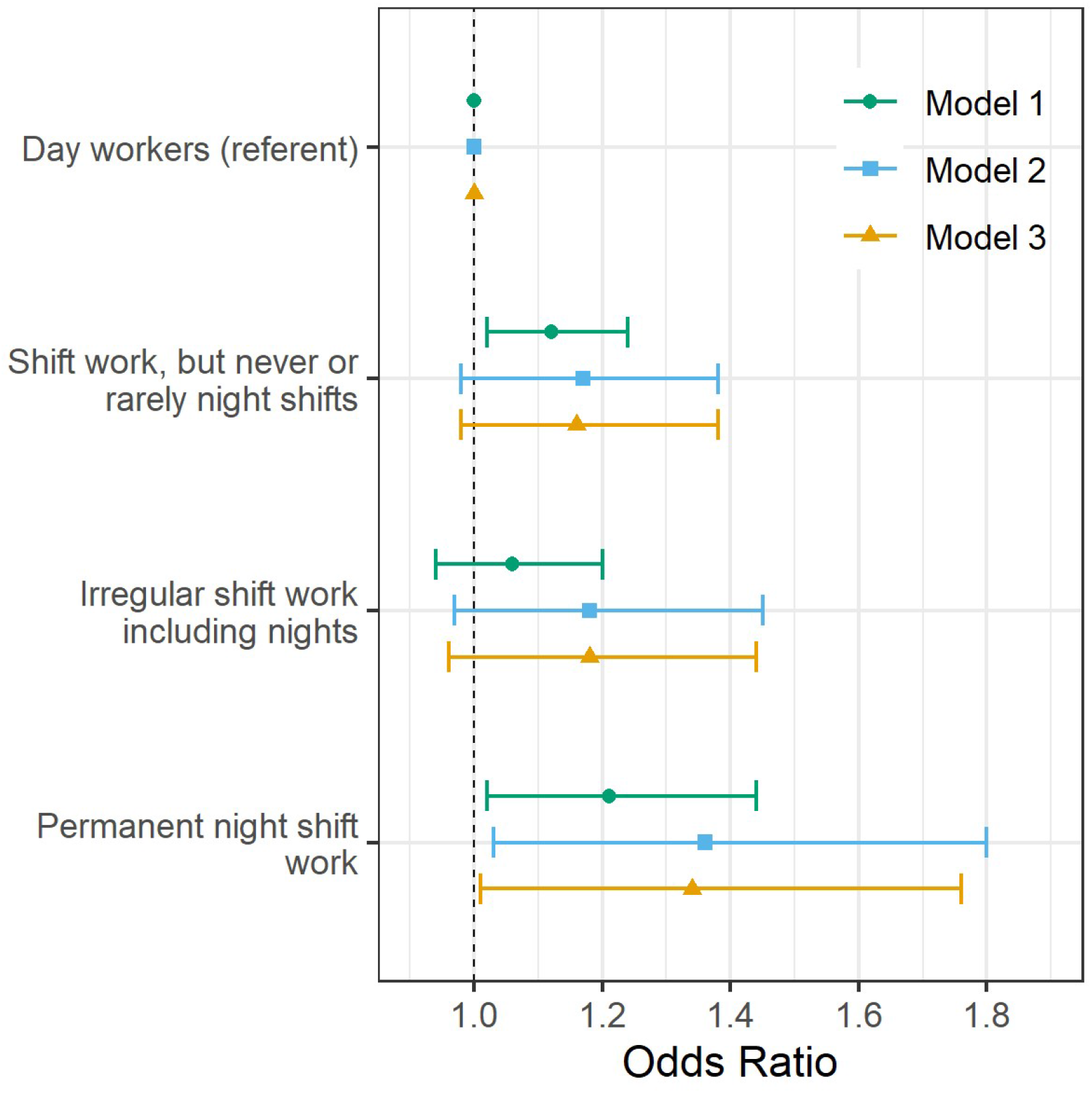
Adjusted odds (95% CI) of moderate-severe asthma by current shift work exposure (N = 257,219). Forest plot of adjusted odds ratios, with corresponding 95% asymptotic confidence intervals, for moderate-severe asthma stratified by current work pattern. Three multivariate logistic regression models were fitted to the data: Model 1 (green circle); age and sex adjusted. Model 2 (blue square) covariates: age, sex, smoking status, smoking pack years, alcohol status, daily alcohol intake, ethnicity, Townsend deprivation index, days exercised (walked, moderate and vigorous), BMI, chronotype, length of working week, job asthma risk and job medical required. Model 3 (yellow triangle); Model 2 covariates plus sleep duration.

A similar pattern of higher odds of asthma was seen when all cases of asthma were considered, **Supplementary Table 1**. In an age- and sex-adjusted model, we observed higher odds of asthma in shift workers who never or rarely worked night shifts when compared to day workers (OR 1.08 (1.02-1.15)). However, this association attenuated to the null with covariate adjustment (model 2). The odds of asthma in shift workers working permanent nights were higher in covariate-adjusted models (Model 2: OR 1.23 (1.03-1.46); model 3: OR 1.20 (1.01-1.43)) than in the age- and sex-adjusted model.

### Symptoms of Asthma

Next we analysed the association between shift work and the experience of wheeze or whistling in the chest in the previous year (N= 280,998). When compared to day workers, the age- and sex-adjusted model revealed higher odds for these symptoms in association with all three types of shift work (shift work, but never or rarely night shifts, irregular night shifts and permanent nights), **Figure 2**. These associations with wheeze or whistling were attenuated but remained significant for all types of shift work in models 2 and 3, (e.g. model 2: shift work, but never or rarely night shifts: OR 1.11 (1.05-1.18); irregular shift work including nights: 1.21 (1.14-1.29); and permanent night shift work: 1.18 (1.08-1.30)).

**Figure 2:**
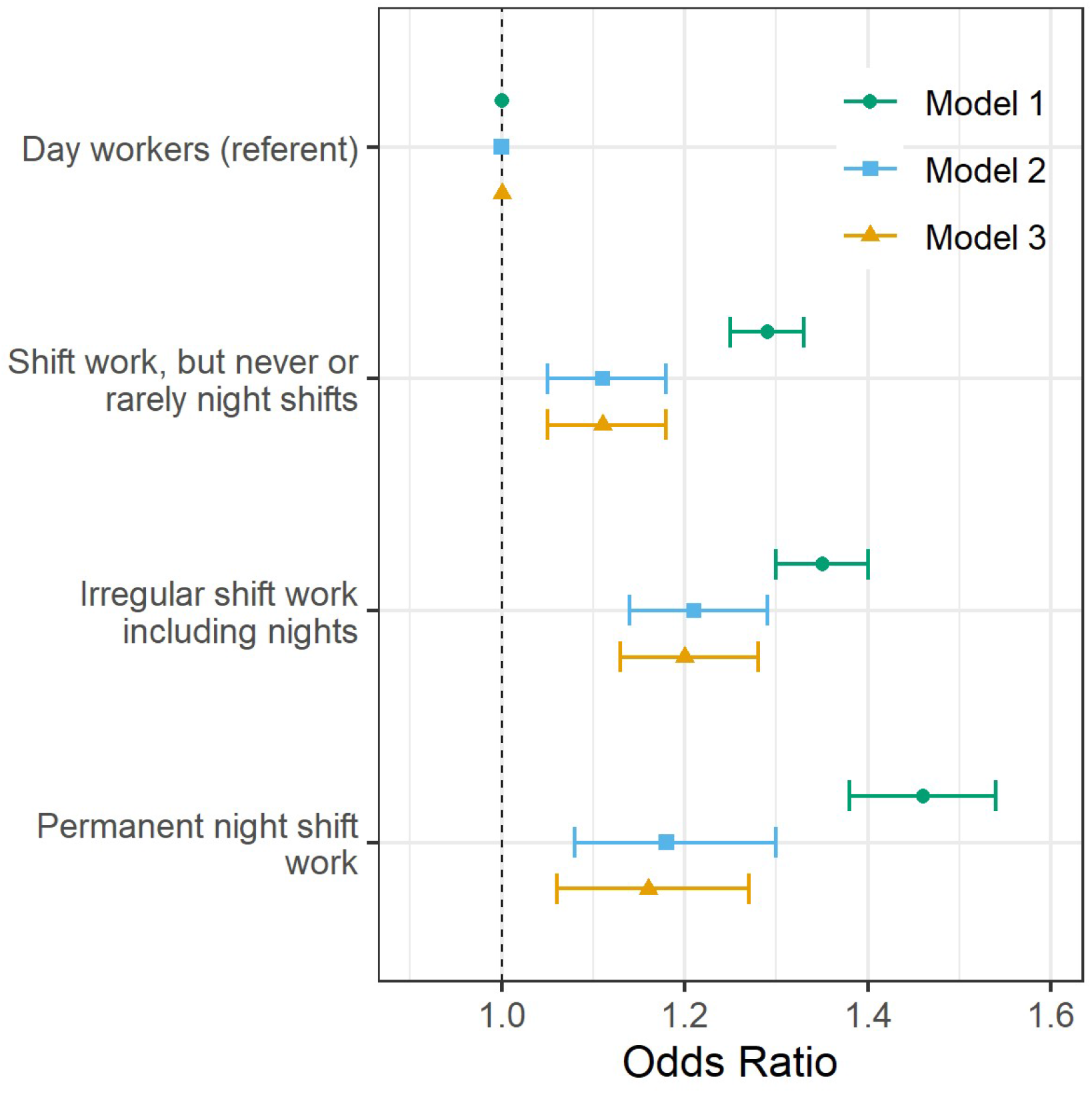
Adjusted odds (95% CI) of experiencing wheeze or whistling in the chest in the last year by current shift work exposure (N = 280,998). Forest plot of adjusted odds ratios, with corresponding 95% asymptotic confidence intervals, for experiencing wheeze or whistling in the chest in the last year stratified by current work pattern. Three multivariate logistic regression models were fitted to the data: Model 1 (green circle); age and sex adjusted. Model 2 (blue square) covariates: age, sex, smoking status, smoking pack years, alcohol status, daily alcohol intake, ethnicity, Townsend deprivation index, days exercised (walked, moderate and vigorous), BMI, chronotype, length of working week, job asthma risk and job medical required. Model 3 (yellow triangle); Model 2 covariates plus sleep duration.

### Obstructive Spirometry

We also examined the association between shift work status and obstructive lung function assessed as the proportion of participants with a forced expiratory volume in 1 second (FEV_1_) that was < 80% of the predicted value based on height and age (N=89,157) (28). In age- and sex-adjusted models there were higher odds of participants having an obstructive FEV_1_ (<80% predicted) in all shift work groups, when compared to day workers, **Table 2**. After multivariable adjustment, these associations attenuated towards the null with higher odds remaining for shift workers who never or rarely worked night shifts and for those working permanent nights (e.g. model 2: shift work, but never or rarely night shifts: OR 1.19 (1.08-1.32); and permanent night shift work: OR 1.20 (1.03-1.41) compared to day workers).

**Table 2:** Adjusted odds (95% CI) of having a critical FEV1 predicted percentage (<80%) by current shift work exposure (N = 89,157)

|  | Current work schedule |  |  |  |
| --- | --- | --- | --- | --- |
|  | Day workers | Shift work, but never or rarely night shifts | Irregular shift work including nights | Permanent night shift work |
| Total cases (% of total sample size) | 9,381 (12.73%) | 1,183 (15.84%) | 849 (15.07%) | 397 (16.99%) |
| Total sample size | 73,719 | 7,469 | 5,632 | 2,337 |
| Model 1: Age and Sex adjusted OR (95% CI) | 1 (referent) | 1.31 (1.23-1.40) | 1.27 (1.18-1.37) | 1.44 (1.29-1.61) |
| Model 2: Multivariable adjusted OR (95% CI) | 1 (referent) | 1.19 (1.08-1.32) | 1.08 (0.96-1.21) | 1.20 (1.03-1.41) |
| Model 3: Model 2 covariates + Sleep Duration (95% CI) | 1 (referent) | 1.19 (1.08-1.32) | 1.08 (0.97-1.21) | 1.21 (1.04-1.42) |

The predicted FEV1 was estimated based on height and age. Model 2 covariates: age, sex, smoking status, smoking pack years, alcohol status, daily alcohol intake, ethnicity, Townsend deprivation index, days exercised (walked, moderate and vigorous), BMI, chronotype, length of working week, job asthma risk and job medical required. Model 3 data are adjusted for Model 2 covariates plus sleep duration.

### Lifetime duration of night shift work

Next we used data on 107,930 participants who provided lifetime work history data. When compared to those reporting no history of shift work, the highest odds for moderate-severe asthma was seen in participants reporting < 5years of shift work (OR (95% CI): 1.34 (1.08-1.66) and the lowest odds when performing ≥ 10 years of shift work (1.22 (1.08-1.38)) **Figure 3a**. In participants reporting < 5 years of shift work, high point estimates for odds for moderate-severe asthma remained after adjusting for covariates in models 2 and 3 but relationships were attenuated to the null for those with higher lifetime durations. No strong statistical evidence of a trend was found when treating lifetime duration of shift work as a continuous variable in any model.

**Figure 3:**
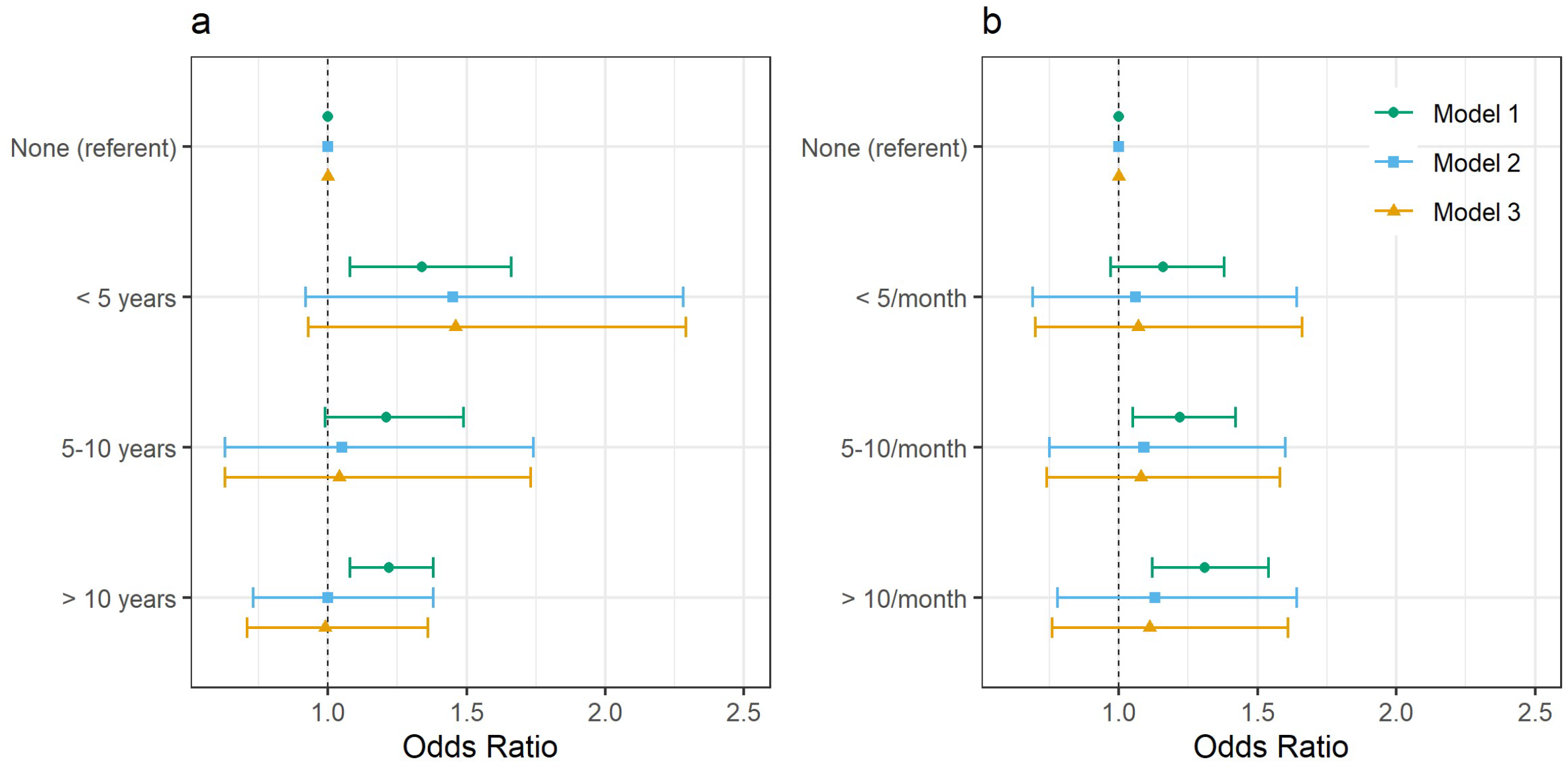
Adjusted odds (95% CI) of moderate-severe asthma by lifetime duration of shift work including nights (a) and by average monthly frequency of shifts that included night shifts (b) (N = 107,930). Forest plot of adjusted odds ratios, with corresponding 95% asymptotic confidence intervals, for moderate-severe asthma stratified by lifetime duration of shift work including nights **(a)** and by average monthly frequency of shifts that included nights **(b)**. Three multivariate logistic regression models were fitted to the data: Model 1 (green circle); age and sex adjusted. Model 2 (blue square) covariates: age, sex, smoking status, smoking pack years, alcohol status, daily alcohol intake, ethnicity, Townsend deprivation index, days exercised (walked, moderate and vigorous), BMI, chronotype, length of working week, job asthma risk and job medical required. Model 3 (yellow triangle); Model 2 covariates plus sleep duration.

### Average lifetime night shift frequency

Using the same historical lifetime work data, we analysed the prior frequency of night shift work in relation to the prevalence of moderate-severe asthma (N=107,930), **Figure 3b**. In age- and sex-adjusted models, when compared to participants reporting no shift work, there were higher odds of moderate-severe asthma in people reporting prior higher frequencies of night shift work (5-10 night shifts/month: (OR (95%CI): 1.22 (1.05-1.42) and also ≥ 10night shifts/month (1.31 (1.12-1.54)), but not the lower frequency of shift work (<5night shifts/month (1.16 (0.97-1.38)). However, these associations attenuated to the null on covariate adjustment (models 2 and 3). There was no strong statistical evidence of average lifetime frequency of night shift work as a continuous variable associating with asthma in any model.

### Chronotype

Chronotype might influence the health impacts of shift work and particularly extreme chronotypes may induce circadian misalignment with the external world, especially in shift workers (23, 24), and so we assessed the relationships between chronotype and asthma in UK Biobank data. Initially, we analysed the likelihood of asthma by chronotype in all UK Biobank participants (N=413,040), **Table 3**. People reporting either extreme chronotype (definitely a morning person or definitely an evening person) had higher odds of having any asthma when compared to those describing themselves as intermediate chronotypes. After adjustment for covariates, the odds ratios for asthma in those reporting definitely a morning person were 1.12 (1.03-1.21) and for those reporting definitely an evening person were 1.16 (1.04-1.28) (model 3).

**Table 3:** Adjusted odds (95% CI) of any asthma by chronotype (N = 413,040)

|  | Chronotype |  |  |
| --- | --- | --- | --- |
|  | Intermediate chronotype | Definitely a morning person | Definitely an evening person |
| Total cases (% of total sample size) | 15,010 (5.68%) | 6,786 (6.06%) | 2,596 (7.06%) |
| Total sample size | 264,279 | 112,007 | 36,754 |
| Model 1: Age and Sex adjusted OR (95% CI) | 1 (referent) | 1.07 (1.04-1.10) | 1.27 (1.22-1.33) |
| Model 2: Multivariable adjusted OR (95% CI) | 1 (referent) | 1.12 (1.04-1.22) | 1.16 (1.05-1.29) |
| Model 3: Model 2 covariates + Sleep Duration (95% CI) | 1 (referent) | 1.12 (1.03-1.21) | 1.16 (1.04-1.28) |

Model 2 covariates: age, sex, smoking status, smoking pack years, alcohol status, daily alcohol intake, ethnicity, Townsend deprivation index, days exercised (walked, moderate and vigorous), BMI, length of working week, job asthma risk and job medical required. Model 3 data are adjusted for Model 2 covariates plus sleep duration.

When we assessed the likelihood of moderate-severe asthma in relation to chronotype (N= 398,252), age- and sex-adjusted models demonstrated higher odds for moderate-severe asthma for people with either extreme chronotype when compared to people with intermediate chronotypes, **Supplementary Table 2**. Results for definitely an evening person attenuated to the null after covariate adjustment (e.g. model 3: OR 1.17 (0.99-1.38)); attenuation was less for definitely a morning person (e.g. model 3: OR 1.19 (1.05-1.35)). Finally, in relation to chronotype, we assessed the likelihood of moderate-severe asthma in individuals with a definite morning chronotype by shift work pattern (N=59,621), **Supplemental Table 3**. In participants who reported being definitely a morning person, there was a higher odds of moderate-severe asthma in covariate-adjusted models in those working irregular shifts, including nights compared to those working day shifts (e.g. model 2: OR 1.55 (1.06-2.27)). There was no excess risk for those morning chronotype workers either on permanent night shifts or rarely working nights.

There was no strong evidence of associations between shift work pattern and the likelihood of moderate-severe asthma when we restricted our analysis to individuals who reported being definitely an evening person (N=20,834) or being an intermediate chronotype (N=148,216), **Supplemental Table 3**. There was no statistical evidence of an interaction between chronotype and shift work in association with asthma (P_interaction_=0.21).

### Asthma Genetic Risk Score

We examined whether genetic susceptibility for asthma modified the relationship between shift work and likelihood of asthma. In those of European ancestry in the UK Biobank cohort, we first showed that higher genetic risk for asthma was associated with a higher odds of moderate-severe asthma (model 2: per risk allele OR 1.13 (1.11-1.16), *P*_*trend*_<0.01, N= 313,816), and for risk of any asthma, (model 2: per risk allele OR 1.12 (1.10-1.13), P_trend_<0.01, N= 302,686). To investigate this effect further, we split the GRS into quartiles and calculated odds of any asthma, **Supplemental Table 4**, and moderate-severe asthma, **Supplemental Table 5**, on these quartiles. Using the quartiles of GRS for moderate-severe asthma, we found that the asthma GRS had a statistically significant interaction on the relationships between odds for moderate-severe asthma and current shift work schedule (p<0.05). However, this interaction did not appear linear in its effects. Odds were higher for moderate-severe asthma in shift workers (who never or rarely worked night shifts) in the second GRS quartile (OR 1.78 (1.17-2.68) and also in permanent night shift workers in the third GRS quartile (OR 2.04 (1.11-3.74), **Supplemental Table 6**.

### Chronic Obstructive Pulmonary Disease (COPD), emphysema and chronic bronchitis

Cases of asthma were defined by including all participants with doctor diagnosed asthma at baseline who were also receiving asthma medication as defined by Shrine et al. 2019 (27). However, this definition may have included participants who had a concurrent doctor diagnosis of COPD, emphysema or chronic bronchitis, since some medications can be used to treat all conditions. There is no way of determining which condition would be predominant amongst these UK Biobank participants, therefore we re-analysed the cohort after excluding all cases of concurrent doctor diagnosed COPD, emphysema and chronic bronchitis. 1790 participants were removed from the any asthma group and 1572 participants from the moderate/severe asthma group. Our results were similar to our previous findings: for moderate/severe asthma, again we found in an age- and sex-adjusted model, there was a higher odds of having moderate-severe asthma in day shift workers who never or rarely undertook night shifts (OR 1.12 (95% CI: 1.01-1.24) when compared to day workers, **Supplemental Table 7**. After adjusting for additional covariates (model 2) only permanent night shift workers had significantly higher likelihood of asthma (OR 1.35 (1.01-1.82)).

Further adjustment for sleep duration slightly attenuated the likelihood of moderate/severe asthma in permanent night shift workers (OR 1.33 (0.99-1.79). In an age- and sex-adjusted model, we observed a higher likelihood of asthma in shift workers who never or rarely worked night shifts when compared to day workers (OR 1.07 (1.01-1.14)). However, this association attenuated to the null after adjusting for additional covariates (model 2). In contrast, the likelihood of asthma in shift workers working permanent nights was statistically significant in multivariable-adjusted models (Model 2: OR 1.26 (1.05-1.5); model 3: OR 1.23 (1.03-1.48)), **Supplemental Table 8**.

## Discussion

We now show that when compared to day workers: a) people working permanent nights had higher adjusted odds of moderate-severe asthma; b) people doing any type of shift work had higher adjusted odds of wheeze or whistling in the chest; c) shift workers who never or rarely worked on nights and people working permanent nights had higher adjusted likelihood of having obstructive spirometry (FEV1 <80% predicted). We analysed data from more than 280,000 UK Biobank participants, 17% of whom were shift workers, which is similar to the reported prevalence of shift work in other industrialized counties (11).

Rotational shift work disrupts the entrainment of endogenous circadian rhythms to external cues in the environment, resulting in circadian misalignment (29, 30). Shift workers, especially those working night shifts, sleep at inappropriate circadian phase, causing circadian misalignment between their sleep-wake behaviour and endogenous circadian processes. Mouse models of shift work have revealed that it is the phase misalignment between the internal clockwork and behaviour that drives many of the resulting pathologies, including metabolic and cardiovascular dysfunction (31). To date, the association between asthma and circadian misalignment has not been investigated. We discover that night shift work associates with an increased risk of asthma.

As the UK Biobank data is drawn from a cross-sectional, observational study, no causal inference is possible. However, it is plausible that circadian misalignment leads to asthma development. To investigate this we looked at people with extreme chronotypes (morning-evening preferences), who experience a degree of circadian alignment in the absence of shift work exposure. Chronotype is genetically determined to some extent (32). We found that extreme chronotypes were significantly more likely to have asthma, even after multivariable adjustments. The majority of individuals in our analysis were day workers (N= 236,897), therefore the higher likelihood of asthma in evening types may represent circadian misalignment caused through conforming to early day shift working hours (33). In fact, evening chronotypes are known to be most at risk for poor health (34). In support of our findings, several previous smaller studies (involving 200 -6000 individuals) have shown that evening chronotype or intermediate chronotype associate with asthma in both adults and adolescents (35, 36). Here, our analysis of chronotype included data from 413,040 individuals including 9604 people with moderate-severe asthma. Furthermore, when we analysed chronotype in the context of type of shift work, we found that there was an increase in moderate/severe asthma risk in morning chronotypes working irregular shifts, including nights (OR 1.55 (1.06-2.27). Morning types find it particularly difficult to adjust to working night shifts (37) and display the highest levels of circadian misalignment. Interestingly, evening chronotypes showed no increase in risk of asthma after shift work exposure, raising the intriguing possibility that evening chronotypes might be protected from the effects of shift work on asthma risk.

We found that the likelihood for any asthma and moderate-severe asthma were higher in individuals working permanent night shifts rather than in those working irregular shift work patterns, including nights. One might assume irregular night shifts lead to more circadian misalignment than permanent night shifts, however only a small minority (<3%) of permanent night shift workers appear to adequately adjust their endogenous circadian timing to night work, as assessed by circadian rhythmicity of melatonin (38). Therefore, we postulate that most shift workers on permanent nights experience a high degree of circadian misalignment; in this case their endogenous circadian rhythms that are important in maintaining autonomic activity (airway tone), immune priming, and stress responses, would be out of alignment with their environment resulting in a susceptibility to developing asthma, increased severity of asthma, or reduced asthma treatment response.

We found a cumulative increase in the odds for moderate-severe asthma in shift workers working more frequent nights; however, this association was attenuated to the null after covariate adjustments and there was no strong statistical evidence when shift work frequency was treated as a continuous variable. We found higher odds of moderate-severe asthma in individuals who had worked night shifts for < 5 years and in those who had worked for ≥ 10 years compared to day workers. The point estimates for the odds ratios suggested that this association might be stronger in individuals working night shifts for < 5 years, compared to those who had worked for ≥ 10 years. We postulate that this might represent the healthy worker effect, where individuals stop working night shifts once their health declines (39). However, these analyses need to be repeated in larger studies.

We devised a GRS for asthma derived from GWAS signals (27) and sought evidence that genetic susceptibility for asthma may modify the risk of shift work exposure. However, the emerging data were inconclusive, with associations being apparent in the middle two quarters of the GRS distribution and not consistent with stronger associations at higher genetic liability as we might have expected. Such an intersection between genetic risk of asthma, and response to shift work exposure would also require replication in a larger cohort.

One intriguing possibility is that rather than night shift work causing asthma people with moderate/severe asthma tend to prefer and self-select for night shift work. This may occur if people with asthma choose to avoid the exacerbation of asthma symptoms during the night by separating in time (rather than summing) the circadian nocturnal trough in lung function (16) from the additional trough in lung function caused by sleep itself (40).

We established that our definition of asthma cases included some individuals with concurrent self-reported doctor-diagnosed COPD, chronic bronchitis or emphysema; the majority of these were present in the moderate/severe asthma group. There is no way to determine in these individuals whether asthma or COPD was the dominant condition from the data within the UK Biobank. Exclusion of these individuals from our analysis did not alter our findings. Past and current smoking is the greatest risk factor for COPD and we took this into account in model 2. It is well-established that there is a degree of overlap between asthma and COPD, particularly in older asthma patients with more fixed airflow obstruction (41). Therefore, we are confident that the association with asthma remains robust even after considering confounding by overlapping chronic inflammatory lung pathologies.

Our study has several strengths; first it involves a large cohort of more than 280,000 individuals from across the UK, with detailed medical history, current employment information, lifestyle information and demographic details, all of which was collected in a uniform manner. Of these >160,000 also had genetic data available. In addition, more than 100,000 individuals from the original cohort also provided detailed employment history.

Our study has some limitations. Firstly, UK Biobank participation rates were low at ∼5%, which may have introduced selection-bias towards more healthy individuals (42). In fact, the overall prevalence of asthma in all participants studied here was ∼5% (also ∼5% in the shift worker cohort alone), compared to ∼10% within the general population of the UK (43). This lower prevalence might also have been influenced by our definition of asthma, which required having both a doctor diagnosis of asthma and currently taking asthma medication (27). This would exclude all those with doctor diagnosed asthma no longer on treatment (childhood asthma), which we felt was appropriate for this study. Furthermore, the UK Biobank data provides no data on younger people and only limited data on ethnic minorities. The sample sizes were small for the morning and evening chronotype analyses, which resulted in low power. There was a reduction in sleep duration reported by night shift workers; this would be a potential confounder and so we took self-reported sleep duration into account in model 3. In fact, we found that model 3 did not significantly alter the results from model 2.

The implications of our research are far-reaching. Approximately 20% of the working population in industrialized countries is involved in some kind of permanent night or rotating shift work (11); we have shown a significant increase in the likelihood of asthma in shift workers working permanent nights. Since there is a high background prevalence of asthma, around 10% of the general population (43), it follows that the prevalence of asthma in shift workers may be even higher. However, there are no specific national clinical guidelines for how to manage asthma in shift workers. Future, prospective clinical studies are required to inform public health policy.

In conclusion, our study has determined that there is an increased likelihood of asthma (especially moderate-severe asthma) in shift workers on permanent nights. This suggests a causal pathway from circadian misalignment to development, or progression of asthma. Modifying shift work schedules to take into account chronotype might present a public health measure to reduce the risk of developing inflammatory diseases, such as asthma.

## Online Methods

### UK Biobank

The UK Biobank study was approved by the National Health Service National Research Ethics Service (ref. 11/NW/0382), and all participants provided written informed consent to participate in the UK Biobank study.

### Shift Work Assessment

We defined shift work as previously reported by Vetter et al (4), however, we combined ‘irregular or rotating shifts with some night shifts’ and ‘irregular or rotating shifts with ususal night shifts’ to form one group ‘irregular shift work including nights’. Briefly, participants employed at baseline were asked to report whether their current main job involved shift work (i.e. a schedule falling outside of 9:00am to 5:00pm; by definition, such schedules involved afternoon, eveninig or night shifts (or rotating though these shifts). If yes, participants were further asked whether their main job involved night shifts, defined as ‘..a work schedule that involves working thoughthe normal sleeping hours, for instance, working though the hours from 12:00am to 6:00am’. For both questions, response options were ‘never/rarely’, ‘sometimes’, ‘usually’, or ‘always’ and included additional options: ‘prefer not to answer’ and ‘do not know’. Based on those two questions, we derived participants’ current shift work status, categorized as ‘day workers’, ‘shift worker, but only rarely if ever nights’, ‘irregular shift work including nights’ and ‘permanent night shifts’.

In the lifetime employment assessment, individuals reported each job ever worked, the number of years in each job ever worked, the number of years in each job, and the number of night shifts per month each job entailed. We restricted our analysis to those individuals who provided in depth lifetime employment information (N= 107,930), we restricted the employment history to only jobs worked prior to 2008, since this was when the diagnosis of asthma was taken at baseline. We aggregated duration (i.e., number of years working night shifts) and frequency (i.e., the average number of night shifts per month) of night shift work.

### Asthma Definition

Cases of asthma were defined by including all those participants with doctor diagnosed asthma at baseline as well as also being on any medication used to treat asthma as defined by Shrine et al. 2019 (27). Cases of moderate-severe asthma were defined as having doctor diagnosed asthma at baseline as well as meeting BTS step 3-5 criteria, i.e. for stage 3 taking β2 agonists plus inhaled corticosteroid; stage 4 taking higher dose inhaled corticosteroids than stage 3 patients and addition of a fourth drug (eg, leukotriene receptor antagonist, theophylline); and stage 5, taking oral corticosteroid or omalizumab, or both (27). We excluded participants with doctor-diagnosed asthma who reported not to be on asthma medication (N=18,806) and those on asthma medication but who did not have doctor diagnosed asthma (N=1,345) from our analyses. When analysing the risk of moderate-severe we further excluded participants with asthma taking medication on BTS stage 1 and 2 (N=9,455).

Within the parameters from the UK Biobank assessment centre data was the question relating to whether a participant had experienced ‘Wheeze or whistling in the chest in the last year’. We excluded participants who answered “Do not know” or “Prefer not to answer” from any statistical analyses. Forced expiratory volume in 1-second (FEV_1_), predicted percentage, was also analysed. FEV_1_ predicted percentages were calculated (44). FEV_1_ predicted percentages were filtered to produce two sub-populations; FEV_1_ ≥ 80% and FEV_1_ < 80%, with the latter indicative of an obstructive respiratory pathology (45, 46) e.g. asthma (47, 48). Participants were split into ‘yes’ and ‘no’ sub-populations for ‘Wheeze or whistling in the chest in the last year’. These and the FEV_1_ predicted percentage sub-populations were further split according to participant’s current work shift schedule, previously outlined.

### Occupational Asthma

We identified participants who were employed in jobs that might lead to the development of occupational asthma. These jobs included bakers, food processors, forestry workers, chemical workers, plastics and rubber workers, metal workers, welders, textile workers, electrical and electronic production workers, storage workers, farm workers, waiters, cleaners, painters, dental workers and laboratory technicians (49-52). We also identified occupations, in which a medical assessment might select against a person with asthma (Protective Service Officers (officers in armed forces, police officers (inspectors and above) and senior officers in fire, ambulance, prison and related services), science technicians and researchers, probation officers and Transport Associate Professionals (including airline pilots and flight engineers, ship and hovercraft officers, train drivers). Both of these were included as covariates in models 2 and 3.

### Chronotype

Participants self-reported chronotype on a touch-screen questionnaire at baseline by answering a question taken from the Morningness-Eveningness questionnaire (question 19;[53]). The question asks: “Do you consider yourself to be….” with response options “Definitely a ‘morning’ person”, “More a ‘morning’ than ‘evening’ person”, “More an ‘evening’ than a ‘morning’ person,” “Definitely an ‘evening’ person,” “Do not know,” and “Prefer not to answer.” Subjects who responded “Do not know” or “Prefer not to answer” were set to missing. This single item has been shown to correlate with sleep timing and dim-light melatonin in set (54-56). For our analyses we combined “more a ‘morning’ than ‘evening’ person” with “more an ‘evening’ than ‘morning’ person” to form an intermediate group. In our initial analysis of chronotype in asthma, we included all individuals with asthma and chronotype information, N= 413,040 (N=398,252 for moderate-severe asthma). Subsequently we investigated shift work in asthma stratified by chronotype (N = 228,671); this excluded participants not in paid employment or self-employed at baseline, or answered “Do not know” or “Prefer not to say” when asked (N=169,581).

### Genetic Risk Score for Asthma

Genotyping in the UK Biobank was performed on two arrays, UK BiLEVE and UK Biobank Aziom. Genotyping, quality control, and imputation procedures have been previously described (57). A total of 488,232 participants in the UK Biobank were genotyped. In total, 337,409 unrelated samples of European ancestry were then filtered and those with an incomplete diagnosis of asthma were excluded, leaving 313,816 for analysis (302,686 for moderate/severe asthma).

We derived a genetic risk score (GRS) for asthma and moderate/severe asthma using 24 GWAS SNPs previously reported by Shrine et al. 2019 (27) for each individual participant. The GRS war generated using PLINK by summing the number of risk (asthma-increasing) alleles, which were weighted by the respective allelic effect size (β-coefficient) from the discovery GWAS. For variants not available in UK Biobank, we used the corresponding proxy SNP as indicated in Table 2 within (27). Scaling of the individual GRS was performed to allow interpretation of the effects as a per-1 risk allele increase in the GRS (division by twice the sum of the β-coefficients and multiplication by twice the square of the SNP count representing the maximum number of risk alleles). Analysis of GRS was performed by subdividing into quartiles, as well as the impact per-1 risk allele. Analysis of the shift work effect on asthma was performed on all GRS quartiles. The interaction between GRS quartiles and shift work schedule was tested and a P value for interaction was computed.

### Statistical Analysis

We fitted a multivariate logistic regression model to the data and used this to estimate adjusted odds ratios and 95% asymptotic confidence intervals on those odds ratios.

In model 1 we initially adjust for participant age and sex. We extend this in model 2 to additionally include BMI, ethnicity, chronotype, Townsend Deprevation Index (TDI), days exercised (walking, moderate exercise and vigorous exercise), smoker status (current, previous or never) and pack years smoked, alcohol status (current, previous or never) and alcohol weekly intake, length of working week and whether current job is considered to have an occupational asthma risk or requires a medical examination prior to hiring. These covariates were chosen by consideration of participant characteristics (**Table 1**). Lastly model 3 also included sleep duration in addition to covariates in model 2 (58).

When investigating continuous variables (lifetime duration and frequency of shift work including nights (**Figure 3**), and odds by genetic risk score (**Supplementary Tables 5 and 6**) p-values for the linear trend were obtained by considering the variable as continuous and running a Wald test to calculate the significance of the variable in our models.

To analyse the effect of GRS and chronotype on the relationship of current job shift schedule on asthma risk we compared models with and without an interaction term (between job shift schedule and GRS/chronotype). The two models were compared using a likelihood ratio test and a p-value indicating the significance of the interaction computed.

## Data Availability

Please contact corresponding author first author for access to data.

---

**Supplemental Figure 1:**
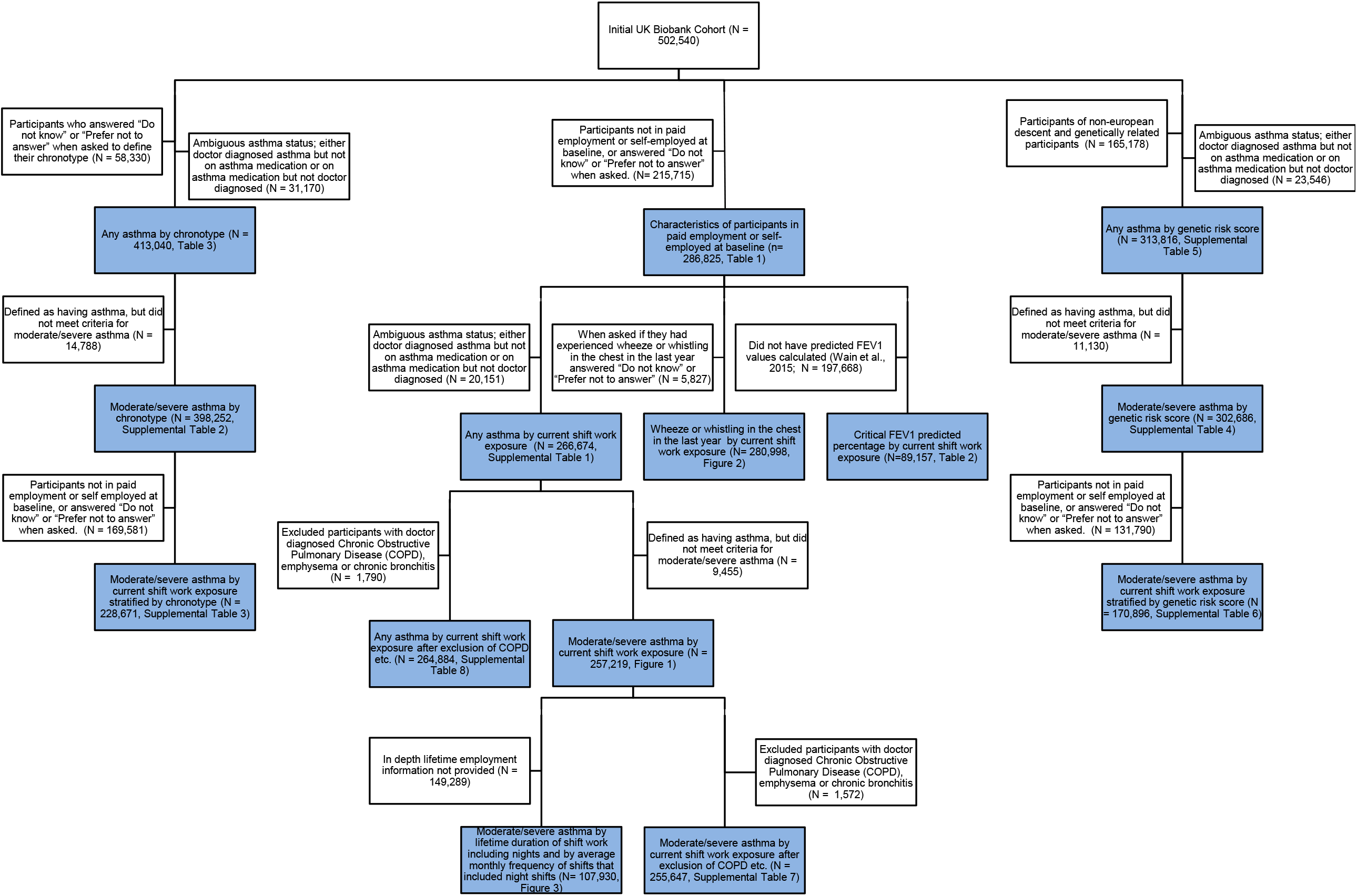
STROBE diagram showing filtering of participants for each analysis. STROBE diagram showing how the full UK Biobank cohort (N=502,540) was filtered for each analysis. Blue boxes correspond to individuals used for the analyses resulting in each figure/table. White boxes show excluded participants at each stage.

**Supplemental Table 1:** Adjusted odds (95% CI) of any asthma by current shift work exposure (N = 266,674)

|  | Current work schedule |  |  |  |
| --- | --- | --- | --- | --- |
|  | Day workers | Shift work, but never or rarely night shifts | Irregular shift work including nights | Permanent night shift work |
| Total cases (% of total sample size) | 11,695 (5.31%) | 1,306 (5.72%) | 872 (5.15%) | 365 (5.48%) |
| Total sample size | 220,234 | 22,838 | 16,945 | 6,657 |
| Model 1: Age and Sex adjusted OR (95% CI) | 1 (referent) | 1.08 (1.02-1.15) | 0.98 (0.91-1.05) | 1.05 (0.95-1.17) |
| Model 2: Multivariable adjusted OR (95% CI) | 1 (referent) | 1.06 (0.95-1.18) | 1.08 (0.95-1.22) | 1.23 (1.03-1.46) |
| Model 3: Model 2 covariates + Sleep Duration (95% CI) | 1 (referent) | 1.06 (0.95-1.18) | 1.07 (0.94-1.21) | 1.20 (1.01-1.43) |

**Supplemental Table 2:**
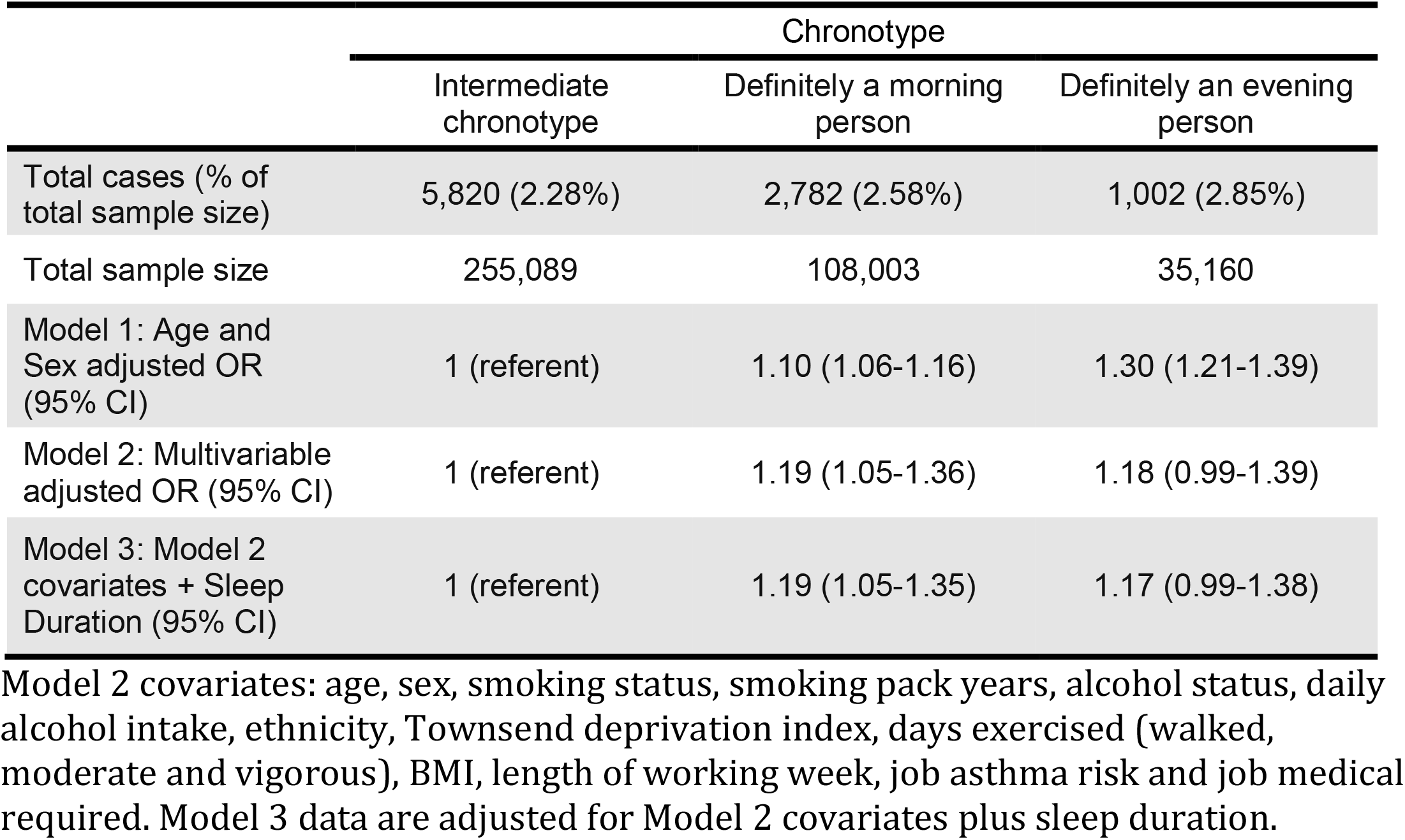
Adjusted odds (95% CI) of moderate-severe asthma by chronotype (N = 398,252)

**Supplemental Table 3:** Adjusted odds (95% CI) and association of moderate-severe asthma and current shift work exposure by chronotype

| Current work schedule | OR (95% CI) | P<sub>interaction</sub> |
| --- | --- | --- |
| Definite morning chronotype (N= 59,621, 1,216 cases) |  |  |
| Day workers | 1 (referent) | 0.21 |
| Shift work, but never or rarely night shifts | 0.97 (0.67-1.39) |  |
| Irregular shift work including nights | 1.55 (1.06-2.27) |  |
| Permanent night shift work | 1.32 (0.69-2.51) |  |
| Intermediate chronotype (N= 148,216, 2,645 cases) |  |  |
| Day workers | 1 (referent) |  |
| Shift work, but never or rarely night shifts | 1.13 (0.90-1.43) |  |
| Irregular shift work including nights | 1.11 (0.84-1.47) |  |
| Permanent night shift work | 1.33 (0.88-2.00) |  |
| Definite evening chronotype (N= 20,834, 447 cases) |  |  |
| Day workers | 1 (referent) |  |
| Shift work, but never or rarely night shifts | 1.18 (0.70-1.99) |  |
| Irregular shift work including nights | 1.10 (0.61-1.99) |  |
| Permanent night shift work | 1.52 (0.88-2.65) |  |

Models were adjusted for covariates in model 2 (age, sex, smoking status, smoking pack years, alcohol status, daily alcohol intake, ethnicity, Townsend deprivation index, days exercised (walked, moderate and vigorous), BMI, length of working week, job asthma risk and job medical required). Interaction p-value is calculated using a LR test comparing the model with and without an interaction term.

**Supplemental Table 4:** Adjusted odds (95% CI) of moderate-severe asthma by genetic risk score (GRS) quartile (N = 302,686)

|  | GRS quartile |  |  |  | p-value<br>for trend |
| --- | --- | --- | --- | --- | --- |
|  | 1st quartile | 2nd quartile | 3rd quartile | 4th quartile |  |
| Total cases (% of total sample size) | 1,166 (1.50%) | 1,585 (2.07%) | 1,906 (2.53%) | 2,707 (3.71%) |  |
| Total sample size | 77,746 | 76,580 | 75,435 | 72,925 |  |
| Model 1: Age and Sex adjusted OR (95% CI) | 1 (referent) | 1.39 (1.29-1.50) | 1.70 (1.58-1.83) | 2.53 (2.36-2.71) | <0.01 |
| Model 2: Multivariable adjusted OR (95% CI) | 1 (referent) | 1.22 (0.99-1.52) | 1.70 (1.39-2.08) | 2.66 (2.21-3.22) | <0.01 |
| Model 3: Model 2 covariates + Sleep Duration (95% CI) | 1 (referent) | 1.23 (0.99-1.52) | 1.71 (1.40-2.09) | 2.67 (2.21-3.23) | <0.01 |

Model 2 covariates: age, sex, smoking status, smoking pack years, alcohol status, daily alcohol intake, Townsend deprivation index, days exercised (walked, moderate and vigorous), BMI, chronotype, length of working week, job asthma risk and job medical required. Model 3 data are adjusted for Model 2 covariates plus sleep duration.

**Supplemental Table 5:** Adjusted odds (95% CI) of any asthma by genetic risk score (GRS) quartile (N = 313,816)

|  | GRS quartile |  |  |  | p-value<br>for trend |
| --- | --- | --- | --- | --- | --- |
|  | 1st quartile | 2nd quartile | 3rd quartile | 4th quartile |  |
| Total cases (% of total sample size) | 3,106 (3.90%) | 3,942 (4.99%) | 4,818 (6.15%) | 6,628 (8.63%) |  |
| Total sample size | 79,686 | 78,937 | 78,347 | 76,846 |  |
| Model 1: Age and Sex adjusted OR | 1 (referent) | 1.30 (1.23-1.36) | 1.62 (1.54-1.69) | 2.33 (2.23-2.43) | <0.01 |
| Model 2: Multivariable adjusted OR (95% CI) | 1 (referent) | 1.24 (1.09-1.41) | 1.52 (1.35-1.72) | 2.33 (2.08-2.61) | <0.01 |
| Model 3: Model 2 covariates + Sleep Duration (95% CI) | 1 (referent) | 1.24 (1.10-1.41) | 1.53 (1.35-1.72) | 2.33 (2.08-2.61) | <0.01 |

Model 2 covariates: age, sex, smoking status, smoking pack years, alcohol status, daily alcohol intake, Townsend deprivation index, days exercised (walked, moderate and vigorous), BMI, chronotype, length of working week, job asthma risk and job medical required. Model 3 data are adjusted for Model 2 covariates plus sleep duration.

**Supplemental Table 6:** Adjusted odds (95% CI) and association of moderate-severe asthma and current shift work exposure by genetic risk

| Current work schedule | OR (95% CI) | P<sub>interaction</sub> |
| --- | --- | --- |
| **GRS first quartile (lowest) (N= 44,088, 475 cases)** |  |  |
| Day workers | 1 (referent) | <0.05 |
| Shift work, but never or rarely night shifts | 1.18 (0.71-1.96) |  |
| Irregular shift work including nights | 0.74 (0.35-1.53) |  |
| Permanent night shift work | 1.67 (0.80-3.51) |  |
| **GRS second quartile (N= 43,396, 667 cases)** |  |  |
| Day workers | 1 (referent) |  |
| Shift work, but never or rarely night shifts | 1.78 (1.17-2.68) |  |
| Irregular shift work including nights | 1.26 (0.72-2.23) |  |
| Permanent night shift work | 1.57 (0.75-3.27) |  |
| **GRS third quartile (N= 42,507, 846 cases)** |  |  |
| Day workers | 1 (referent) |  |
| Shift work, but never or rarely night shifts | 1.41 (0.95-2.09) |  |
| Irregular shift work including nights | 1.22 (0.74-2.02) |  |
| Permanent night shift work | 2.04 (1.11-3.74) |  |
| **GRS fourth quartile (highest) (N= 40,905, 1,218 cases)** |  |  |
| Day workers | 1 (referent) |  |
| Shift work, but never or rarely night shifts | 0.93 (0.64-1.35) |  |
| Irregular shift work including nights | 1.42 (0.97-2.10) |  |
| Permanent night shift work | 1.52 (0.90-2.56) |  |

Models were adjusted for covariates in model 2 (age, sex, smoking status, smoking pack years, alcohol status, daily alcohol intake, ethnicity, Townsend deprivation index, days exercised (walked, moderate and vigorous), BMI, chronotype, length of working week, job asthma risk and job medical required). Interaction p-value is calculated using a LR test comparing the model with and without an interaction term.

**Supplemental Table 7:**
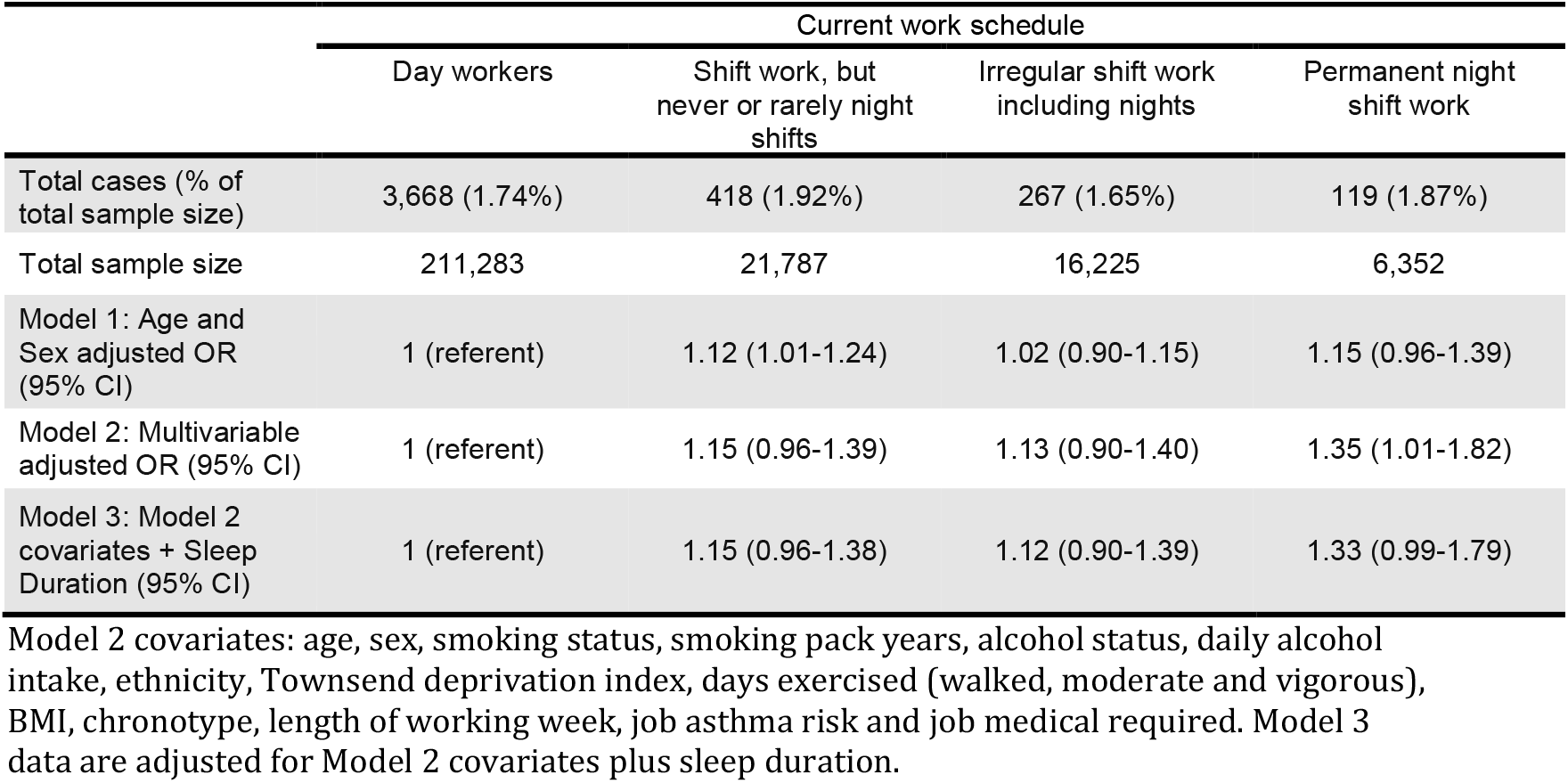
Adjusted odds (95% CI) of moderate-severe asthma by current shift work exposure after excluding participants with doctor diagnosed Chronic Obstructive Pulmonary Disease (COPD), emphysema or chronic bronchitis (N = 255,647)

**Supplemental Table 8:** Adjusted odds (95% CI) of any asthma by current shift work exposure after excluding participants with doctor diagnosed Chronic Obstructive Pulmonary Disease (COPD), emphysema or chronic bronchitis (N = 264,884)

|  | Current work schedule |  |  |  |
| --- | --- | --- | --- | --- |
|  | Day workers | Shift work, but never or rarely night shifts | Irregular shift work including nights | Permanent night shift work |
| Total cases (% of total sample size) | 11,290 (5.16%) | 1,247 (5.51%) | 823 (4.90%) | 349 (5.30%) |
| Total sample size | 218,905 | 22,616 | 16,781 | 6,582 |
| Model 1: Age and Sex adjusted OR (95% CI) | 1 (referent) | 1.07 (1.01-1.14) | 0.96 (0.89-1.03) | 1.04 (0.93-1.16) |
| Model 2: Multivariable adjusted OR (95% CI) | 1 (referent) | 1.04 (0.93-1.16) | 1.05 (0.92-1.19) | 1.26 (1.05-1.50) |
| Model 3: Model 2 covariates + Sleep Duration (95% CI) | 1 (referent) | 1.04 (0.93-1.16) | 1.04 (0.91-1.18) | 1.23 (1.03-1.48) |

## References

1. Michael H. Smolensky, Ramon C. Hermida, Alain Reinberg, Linda Sackett-Lundeen & Francesco Portaluppi. Circadian disruption: New clinical perspective of disease pathology and basis for chronotherapeutic intervention, Chronobiology International 2016;33:1101–1119, DOI: 10.1080/07420528.2016.1184678).

2. Sun M, Feng W, Wang F, Zhang L, Wu Z, Li Z, Zhang B, He Y, Xie S, Li M, Fok JPC, Tse G, Wong MCS, Tang JL, Wong SYS, Vlaanderen J, Evans G, Vermeulen R, Tse LA. Night shift work exposure profile and obesity: Baseline results from a Chinese night shift worker cohort. PLoS One. 2018;13(5):e0196989. Published 2018 May 15.doi:10.1371/journal.pone.0196989

3. Zimberg IZ, Fernandes Junior SA, Crispim CA, Tufik S, de Mello MT. Metabolic impact of shift work. Work. 2012;41 Suppl 1:4376–83

4. Vetter C, Dashti HS, Lane JM, et al. Night Shift Work, Genetic Risk, and Type 2 Diabetes in the UK Biobank. Diabetes Care. 2018;41(4):762–769. doi:10.2337/dc17-1933

5. Jankowiak S, Backé E, Liebers F, et al. Current and cumulative night shift work and subclinical atherosclerosis: results of the Gutenberg Health Study. Int Arch Occup Environ Health. 2016;89(8):1169–1182. doi:10.1007/s00420-016-1150-6

6. Wegrzyn LR, Tamimi RM, Rosner BA, et al. Rotating Night-Shift Work and the Risk of Breast Cancer in the Nurses’ Health Studies. Am J Epidemiol. 2017;186(5):532–540. doi:10.1093/aje/kwx140

7. Papantoniou K, Devore EE, Massa J, et al. Rotating night shift work and colorectal cancer risk in the nurses’ health studies. Int J Cancer. 2018;143(11):2709–2717. doi:10.1002/ijc.31655

8. Mason, I.C., Qian, J., Adler, G.K. et al. Impact of circadian disruption on glucose metabolism: implications for type 2 diabetes. Diabetologia 63, 462–472 (2020). https://doi.org/10.1007/s00125-019-05059-6

9. Chellappa SL, Vujovic N, Williams JS, Scheer FAJL. Impact of Circadian Disruption on Cardiovascular Function and Disease. Trends Endocrinol Metab. 2019 Oct;30(10):767–779. doi: 10.1016/j.tem.2019.07.008. Epub 2019 Aug 16.

10. Castanon-Cervantes OL, Wu M, Ehlen JC, Paul K, Gamble KL, Johnson RL, Besing RC, Menaker M, Gewirtz AT, Davidson AJ. Dysregulation of inflammatory responses by chronic circadian disruption. J Immunol. 2010 Nov 15;185(10):5796–805. doi: 10.4049/jimmunol.1001026. Epub 2010 Oct 13.

11. Haus, E. & Smolensky, M. Biological Clocks and Shift Work: Circadian Dysregulation and Potential Long-term Effects. Cancer Causes Control 2006;17: 489. https://doi.org/10.1007/s10552-005-9015-4

12. The Global Asthma Report 2018. Auckland, New Zealand: Global Asthma Network, 2018

13. Mukherjee M, Stoddart A, Gupta RP, et al. The epidemiology, healthcare and societal burden and costs of asthma in the UK and its member nations: analyses of standalone and linked national databases. BMC Med. 2016;14(1):113. Published 2016 Aug 29. doi:10.1186/s12916-016-0657-8

14. Nurmagambetov T, Kuwahara R, Garbe P. The Economic Burden of Asthma in the United States, 2008 – 2013. Annals of The American Thoracic Society 2017.

15. Turner-Warwick M. Nocturnal asthma: a study in general practice. J R Coll Gen Pract 1989;39:239–243.

16. Sutherland ER. Nocturnal asthma. J Allergy Clin Immunol 2005;116:1179–1186, quiz 1187.

17. Durrington HJ, Gioan-Tavernier GO, Maidstone RJ, Krakowiak K, Loudon ASI, Blaikley JF, Fowler SJ, Singh D, Simpson A, Ray DW. Time of Day Affects Eosinophil Biomarkers in Asthma: Implications for Diagnosis and treatment. Am J Respir Crit Care Med. 2018 Aug 29. doi: 10.1164/rccm.201807-1289LE. [Epub ahead of print]

18. Spengler CM, Shea SA. Endogenous circadian rhythm of pulmonary function in healthy humans. Am J Respir Crit Care Med. 2000 Sep;162(3 Pt 1):1038–46.

19. Attarchi, M., Dehghan, F., Yazdanparast, T., Mohammadi, S., Golchin, M., Sadeghi, Z., Moafi, M., & Seyed Mehdi, S. M. (2014). Occupational asthma in a cable manufacturing company. Iranian Red Crescent medical journal, 16(10), e9105. https://doi.org/10.5812/ircmj.9105)

20. L. J. Palmer UK Biobank: Bank on it. Lancet 369, 1980–1982 (2007). doi:10.1016/S0140-6736(07)60924-6 pmid:17574079

21. Fischer D, Lombardi DA, Marucci-Wellman H, Roenneberg T. Chronotypes in the US - Influence of age and sex. PLoS One. 2017 Jun 21;12(6):e0178782. doi: 10.1371/journal.pone.0178782. eCollection 2017.

22. Paine SJ, Gander PH, Travier N. The epidemiology of morningness/eveningness: influence of age, gender, ethnicity, and socioeconomic factors in adults (30-49 years). J Biol Rhythms. 2006 Feb;21(1):68–76.

23. Juda, M., Vetter, C., & Roenneberg, T. (2013). Chronotype Modulates Sleep Duration, Sleep Quality, and Social Jet Lag in Shift-Workers. Journal of Biological Rhythms, 28(2), 141–151).

24. Vetter C, Fischer D, Matera JL, Roenneberg T. Aligning shift times with chronotype improves sleep and reduces circadian disruption. Curr Biol. 2015 Mar 30;25(7):907–11. doi: 10.1016/j.cub.2015.01.064. Epub 2015 Mar 12.

25. Vicente CT, Revez JA, Ferreira MAR. Lessons from ten years of genome-wide association studies of asthma. Clin Transl Immunology 2017;6(12):e165

26. Allen NE, Sudlow C, Peakman T, Collins R, on behalf of UK Biobank. UK Biobank Data: Come and Get It. Science Translational Medicine19 Feb 2014: 224ed4

27. Shrine N, Portelli MA, John C, Soler Artigas M, Bennett N, Hall R, Lewis J, Henry AP, Billington CK, Ahmad A, Packer RJ, Shaw D, Pogson ZEK, Fogarty A, McKeever TM, Singapuri A, Heaney LG, Mansur AH, Chaudhuri R, Thomson NC, Holloway JW, Lockett GA, Howarth PH, Djukanovic R, Hankinson J, Niven R, Simpson A, Chung KF, Sterk PJ, Blakey JD, Adcock IM, Hu S, Guo Y, Obeidat M, Sin DD, van den Berge M, Nickle DC, Bossé Y, Tobin MD, Hall IP, Brightling CE, Wain LV, Sayers I. Moderate-to-severe asthma in individuals of European ancestry: a genome-wide association study. Lancet Respir Med. 2019 Jan;7(1):20–34. doi: 10.1016/S2213-2600(18)30389-8. Epub 2018 Dec 11.

28. Wain LV, Shrine N, Miller S, Jackson VE, Ntalla I, Soler Artigas M, Billington CK, et al. Novel insights into the genetics of smoking behaviour, lung function, and chronic obstructive pulmonary disease (UK BiLEVE): a genetic association study in UK Biobank. Lancet Respir Med. 2015 Oct;3(10):769–81. doi: 10.1016/S2213-2600(15)00283-0. Epub 2015 Sep 27.

29. Fischer D, Vetter C and Roenneberg T. A novel method to visualise and quantify circadian misalignment. Sci Rep. 2016; 6: 38601.Published online 2016 Dec 8. doi: 10.1038/srep38601

30. Sletten TL, Cappuccio FP, Davidson AJ, Van Cauter E, Rajaratnam SMW, Scheer FAJL. Health consequences of circadian disruption. Sleep. 2020 Jan 13;43(1). pii: zsz194. doi: 10.1093/sleep/zsz194.

31. West AC, Smith L, Ray DW, Loudon ASI, Brown TM, Bechtold DA. Misalignment with the external light environment drives metabolic and cardiac dysfunction. Nat Commun. 2017;8(1):417. Published 2017 Sep 12. doi:10.1038/s41467-017-00462-2

32. Jones, S.E., Lane, J.M., Wood, A.R. et al. Genome-wide association analyses of chronotype in 697,828 individuals provides insights into circadian rhythms. Nat Commun 2019;10:343. https://doi.org/10.1038/s41467-018-08259-7)).

33. Abbott SM, Reid KJ, Zee PB. Circadian disorders of the sleep-wake cycle. In: principles and oractice of sleep medicine. 6th Edition. 414-423. Kryger M, Rth T, Dement WC (Eds), Elsevier, Philadelphia.

34. Roeser K, Obergfell F, Meule A, Vogele C, Schlarb AA, Kubler A. Of larks and heartsmorningness/eveningness, heart rate variability and cardiovascular stress response at different times of day. Physiol Behav 2012;106:151–157

35. Ferraz E, Borges MC, Vianna EO. Influence of nocturnal asthma on chronotype. J. Asthma 2008;45:911–915

36. Haldar P, Debnath S, Maity SG, Mitra R, Biswas M, Bhattacharjee S, Saha S, Das A, Sengupta S, Annesi-Maesano I, Garcia-Aymerich J, Ghosh A, Bandyopadhyay A, Chattopadhyay D, Moitra S and Moitra S. Association between asthma and allergic diseases and circadian preference (chronotype) of the adolescents. ERJ 2019;54:PA2783

37. Gamble KL, Motsinger-Reif AA, Hida A, Borsetti HM, Servick SV, Ciarleglio CM, Robbins S, Hicks J, carver K, Hamilton N et al. Shift work in Nurses: Contribution of Phenotypes and Genotypes to Adaption. Plos One 2011; 6:e18395

38. Folkard S. Do permanent night workers show circadian adjustment? A review based on the endogenous melatonin rhythm. Chronobiology International 2008;25:215–224

39. Knutsson A, Åkerstedt T. The healthy-worker effect: Self-selection among Swedish shift workers. Work Stress. 1992;6:163–7.

40. Robert D. Ballard (1999) Sleep, Respiratory Physiology, and Nocturnal asthma, Chronobiology International, 16:5, 565–580, DOI: 10.3109/07420529908998729

41. http://www.ginasthma.org Global Initiative for Asthma: Diagnosis of Diseases of Chronic Airflow Limitation: Asthma, COPD and Asthma-COPD Overlap Syndrome (ACOS)

42. Fry A, Littlejohns TJ, Sudlow C, Doherty N, Adamska L, Sprosen T, Collins R, Allen NE. Comparison of Sociodemographic and Health-Related Characteristics of UK Biobank Participants With Those of the General Population. Am J of Epidem 2017;186:1026–1034, https://doi.org/10.1093/aje/kwx246

43. Asthma UK. www.asthma.org.uk

44. Wain LV, Shrine N, Miller S et al. Novel insights into the genetics of smoking behaviour, lung function, and chronic obstructive pulmonary disease (UK BiLEVE): a genetic association study in UK Biobank. Lancet Respir Med. 2015; 3: 769–781

45. Gauderman, W. J., Avol, E., Gilliland, F., Vora, H., Thomas, D., Berhane, K., McConnell, R., Kuenzli, N., Lurmann, F. & Rappaport, E. (2004). ‘The effect of air pollution on lung development from 10 to 18 years of age’, New England Journal of Medicine, 351(11), pp. 1057–1067.

46. Criner, G. J., Martinez, F. J., Anzueto, A., Barnes, P. J. & Bourbeau, J. (2017). ‘Global Strategy for the Diagnosis, Management and Prevention of Chronic Obstructive Lung Disease 2017 Report’.

47. Global Initiative for Asthma, P. (2017). ‘Global Strategy for Asthma Management and Prevention: 2017 Update’.

48. Cukic, V., Lovre, V., Dragisic, D. & Ustamujic, A. (2012). ‘Asthma and chronic obstructive pulmonary disease (COPD)–differences and similarities’, Materia socio-medica, 24(2), p. 100.46. BTS/SIGN British Guideline on the management of asthma 2019

49. Jaakkola JJ, Pilpari R, Jaakkola MS. Occupation and asthma; a population-based incident case-control study. Am J Epidemiol 2003;158:981–7

50. Johnson AR, Dimich-Ward HD, Manfreda J, Becklake MR, Ernst P, Sears MR, Bowie DM, Sweet L, Chan-Yeung M. Occupational asthma in adults in six Canadian communities. Am J Respir Crit Care Med 2000;162:2058–62

51. Kogevinas M, Anto JM, Soriano JB, Tobias A, Burney P. The risk of asthma attributable to occupational exposures. A population-based study in Spain. Spanish Group of the European Asthma Study. Am J Respir Crit Care Med 1996;154:137–43

52. Kogevinas M, Anto JM, Sunyer J, Tobias A, Kromhout H, Burney P. Occupational asthma in Europe and other industrialised areas: a population-based study. European Community Respiratory Health Survey Study Group. Lancet 1999;353:1750–4

53. Horne JA, Ostberg O. A self-assessment questionnaire to determine morningness-eveningness in human circadian rhythms. Int J Chronobiol 1976;4:97–110

54. Kitamura S, Hilda A, Aritake S et al. Validity of the Japanese version of the Munich Chronotype Questionnaire. Chronobiol INt 2014;31:845–850.

55. Megdal SP, Schernhammer ES. Correlates for poor sleepers in a Los Angeles high school. Sleep Med 2007;9:60–63

56. Taillard J, Philip P, Chastang JF, Bioulac B. Validation of Horne and Ostberg morningness eveningness questionnaire in a middle-aged population of French workers. J Biol Rythms 2004;19:76–86

57. Lane JM, Vlasac I, Anderson SG, et al. Genome-wide association analysis identifies novel loci for chronotype in 100,420 individuals from the UK Biobank. Nat Commun 2016;7:10889

58. Koinis-Mitchell D, Kopel SJ, Seifer R, et al. Asthma-related lung function, sleep quality, and sleep duration in urban children. Sleep Health. 2017;3(3):148–156. doi:10.1016/j.sleh.2017.03.008

